# CoViD–19: An Automatic, Semiparametric Estimation Method for the Population Infected in Italy

**DOI:** 10.1101/2020.03.14.20036103

**Authors:** Livio Fenga

## Abstract

To date, official data on the number of people infected with the SARS-CoV-2 - responsible for the CoViD–19 - have been released by the Italian Government just on the basis of a non representative sample of population which tested positive for the swab. However a reliable estimation of the number of infected, including asymptomatic people, turns out to be crucial in the preparation of operational schemes and to estimate the future number of people, who will require, to different extents, medical attentions. In order to overcome the current data shortcoming, this paper proposes a bootstrap–driven, estimation procedure for the number of people infected with the SARS-CoV-2. This method is designed to be robust, automatic and suitable to generate estimations at regional level. Obtained results show that, while official data at March the 12th report 12.839 cases in Italy, people infected wiyh the SARS-CoV-2 could be as high as 105.789.

## 1. Introduction

Cases of COVID-19 break out in Italy where it is first attested a capillary spread of this disease in the European continent after the Asian one: the scenario that is developing in these days is creating an example that unfortunately will certainly be repeated in other states all over the world. In this framework, the availability of a reliable data sources on the diffusion of SARS-CoV-2 – the virus responsible for this disease - is crucial in many ways. It is needed to maximize coordination among emergency services located in different parts of the County and within EU, it is crucial for the preparation of operational schemes, and pivotal to allow a proper prediction of the development of the pandemic.

At the moment, official data on the infection in Italy are based on non random, non representative samples of the population: as a matter of fact people are tested for SARS-CoV-2 on the condition that some symptoms related to the virus are present. These data can ensure a proper estimation of total deaths and total hospitalizations due to the virus-related disease: this is crucial to proceed in terms of optimization available resources, of rationalization of accesses to hospitals, of other health facilities and so forth. Nonetheless, form a pure statistical point of view they are not suitable to provide a reliable source of information on the real number of infected people (thereafter “positive cases”).

Starting from the number of deaths and the number of people tested positive to the virus and improving on the methodology originally proposed by Pueyo (2020), this paper aims to estimate the real number of people infected by the SARS-CoV-2, simply called CORONAVIRUS, in each of the 20 Italian regions.

Small sample size – which is suitable to lead to a strong bias in asymptotic results and which is very likely to imply the construction of incorrect confidence intervals – and the distortion of the sample introduced by the mentioned testing strategy are the two mayor obstacles in reliable estimations.

The presented procedure is designed to overcome these problems. As it will be detailed in the sequel, in order to reduce the impact of biasing components on the parameter estimations, a recent bootstrap scheme, called Maximum Entropy Bootstrap and proposed by Vinod et al. (2009), has been employed. In addition to that, a distance measure – based on the theory of stochastic processes and proposed by Piccolo (1990) – has been employed to guarantee statistical coherence among all the Italian regions.

## 2. The proposed method

In small data sets it is essential to save degrees of freedom (DOF). In this perspective, the adopted model — of the type semiparametric – consists of two parts: a purely non-parametric and a parametric one. While the former does not pose problems in terms of DOF, the latter clearly does. However, the sacrifice in terms of DOF is very limited as an autoregressive model of order 1 (employed in a suitable distance function, as below illustrated) has proved sufficient for the purpose. DOF–saving strategy is also the driving force of the choice not to consider as an exogenous parameter the georeferencing of Regions or to include the regional population in a regression–like scheme but to implicitly assumed these variable embedded in the dynamic of the time series in question.

## 3. Data and contageon indicator

The paper makes use of official data published by Italian Authorities, on the following two variables of interest

1. number of deaths from CoViD–19 (denoted by the Latin letter *M*)
2. number of currently positive cases recorded after the administration of the test (denoted by the Latin letter *C*).

The data set includes 18 daily datapoints collected at regional level during the period of February 24^*th*^ to March 12^*th*^. The total number of Italian regions considered is 20. However, one special administrative area (Trentino Alto Adige) is divided in two subregions, i.e. Trento and Bolzano. Therefore, the set containing all the Italian regions − called Ω – has cardinality |·| = 22 (the cardinality function is denoted by the symbol | · | = 22). Two different subsets are built from Ω i.e. Ω^•^ – containing the regions for which at least one death, out of the group of tested people, has been recorded and Ω^°^ (no recorded deaths):

1. Ω^•^≡ *Piemonte, Lombardia, Veneto, Friuli, Liguria, Emilia, Toscana, Marche, Lazio, Abbruzzo, V alleAosta, Bolzano, Campania, Puglia, Sicilia*
2. Ω^°^ ≡ *Trento, Umbria, Molise, Basilicata, Calabria, Sardegna*,

being Ω ≡ Ω^•^ ∪ Ω^°^. In what follows, the two superscripts ^•^ and ^°^ will be always used respectively with reference to the regions {*r*_1_, *r*_2_, … *r*_15_} *∈* Ω^•^ and in {*s*_1_, *s*_2_, … *s*_6_} ∈ Ω^°^. The time span is denoted as {1, 2, …, *T*}.

In the case of the regions included in Ω^•^, following Pueyo (2020), estimates the total number of people infected by CoViD-19 as follows:

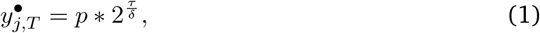

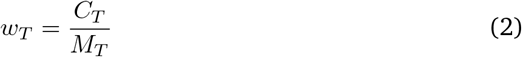

where the superscript • identifies the regions {*r*_1_, *r*_2_, … *r*_15_} ∈ Ω^•^, w is the ratio between current positive cases (C) and number of deaths (M) (2), *τ* the average doubling time for the CoViD–19 (i.e. the average span of time needed for the virus to double the cases) and *δ* the average time for an infected person to die. These two constant terms have been kept fixed as estimated according the data so far available worldwide (see Pueyo (2020)). They are as follows: *τ* = 17.3 and *δ* = 6.2.

The case of the regions belonging to Ω^°^ is more complicated. The approach adopted is as follows:

1. Given the *s*_*j*_ ∈ Ω^°^ a series *c*^*π*^ ∈ Ω^•^ minimizing of a suitable distance function – denoted by the Greek letter *π*(·) – is found. In symbols: 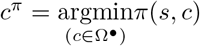;
2. the estimated number of infected at the population level found for *c*^*π*^, say *I*_*c*_^π^ becomes the weight for which the total cases recorded for *s*_*j*_, i.e.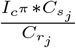 Therefore, the estimate of the variable of interest for this case is as follows:

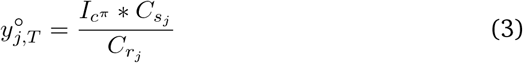

The distance function adopted (*π*), called AR distance, has been introduced by Piccolo (2007)). Briefly, the series of interest are considered a realization of an ARMA (Autoregressive Moving Average) model (see, e.g. Makridakis and Hibon (1997)) so that, each of them can be expressed as an autoregressive model of infinite order, i.e. *AR*(∞) whose infinite sequence of AR parameters is *α*_1_, *α*_2_, ….

Without loss of generality, the distance between the series s and c *π*(*s, c*) (Eqn 3) is expressed as

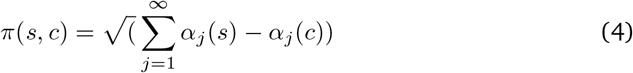

## 4. The Resampling Method

The bootstrap scheme adopted proved to be a real asset for the problem at hand. Given the pivotal role played it will be briefly presented. In essence, the choice of the most appropriate resampling method is far from being an easy task, especially when the identical and independent distribution *iid* assumption (Efron’s initial bootstrap method) is violated. Under dependence structures embedded in the data, simple sampling with replacement has been proved – see, for example Carlstein et al. (1986) – to yield suboptimal results. As a matter of fact, *iid*–based bootstrap schmes are not designed to capture, and therefore replicate, dependence structures. This is especially true under the actual conditions (small sample sizes). In such cases, selecting the “right” resampling scheme becomes a particularly challenging task. Several *ad hoc* methods have been therefore proposed, many of which now freely and publicly available in the form of powerful routines working under software package such as Python^®^ or R^®^. In more details, while in the classic bootstrap an ensemble **Ω** represents the population of reference the observed time series is drawn from, in *MEB* a large number of ensembles (subsets), say { ***ω***_1_, …, ***ω***_*N*_ } becomes the elements belonging to **Ω**, each of them containing a large number of replicates. { *x*_1_, …, *x*_*J*_ } Perhaps, the most important characteristic of the *MEB* algorithm is that its design guarantees the inference process to satisfy the ergodic theorem. Formally, denoting by the symbol | ***·*** | the cardinality function (counting function) of a given ensemble of time series {*x*_*t*_ ∈ ***ω***_*i*_; *i* = 1, …, *N* }, the *MEB* procedure generates a set of disjoint subsets **Ω**_***N***_ ≡ *ω*_1_ ∩ *ω*_1_ ∩ *ω*_*N*_ s.t. 𝔼**Ω**_***N***_ ≈ *µ*(*x*_*t*_), being *µ*(·) the sample mean. Furthermore, basic shape and probabilistic structure (dependency) is guaranteed to be retained 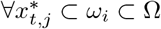.

*MEB* resampling scheme has not negligible advantages over many of the available bootstrap methods: it does not require complicated tune up procedures (unavoidable, for example, in the case of resampling methods of the type Block Bootstrap) and it is effective under non-stationarity. *MEB* method relies on the entropy theory and the related concept of (un)informativeness of a system. In particular, the Maximum Entropy of a given density *δ*(*x*), is chosen so that the expectation of the Shannon Information *H* = 𝔼(*−* log *δ*(*x*)), is maximized, i.e.

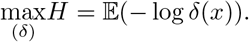

Under mass and mean preserving constraints, this resampling scheme generates an ensemble of time series from a density function satisfying (4). Technically, *MEB* algorithm can be broken down, following Koutris et al. (2008), in 8 steps. They are:

1. a sorting matrix of dimension *T ×* 2, say *S*_1_, accommodates in its first column the time series of interest *x*_*t*_ and an Index Set – i.e. *I*_*ind*_ ={ 2, 3, …, *T*} – in the other one;
2. *S*_1_ is sorted according to the numbers placed in the first column. As a result, the order statistics ***x***_**(*t*)**_ and the vector *I*_*ord*_ of sorted *I*_*ind*_ are generated and respectively placed in the first and second column;
3. compute “intermediate points”, averaging over successive order statistics, i.e.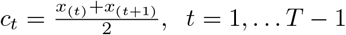, *t* = 1, … *T −* 1 and define intervals *I*_*t*_ constructed on *c*_*t*_ and *r*_*t*_, using *ad hoc* weights obtained by solving the following set of equations:

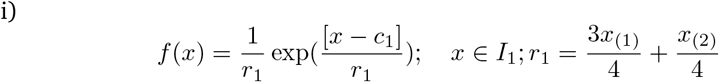

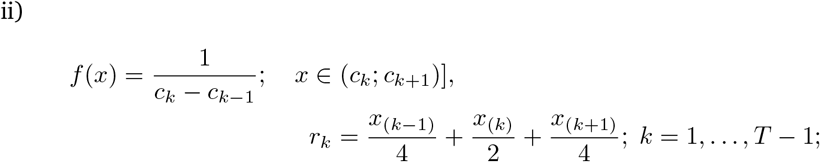

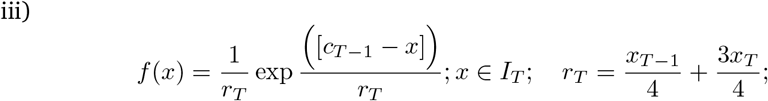
4. from a uniform distribution in [0, 1], generate *T* pseudorandom numbers and define the interval *R*_*t*_ = (*t/T*; *t* + 1*/T*] for *t* = 0, 1, …, *T* − 1, in which each *p*_*j*_ falls;
5. create a matching between *R*_*t*_ and *I*_*t*_ according to the following equations:

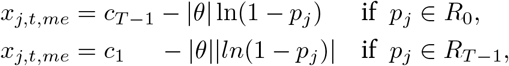

so that a set of *T* values { *x*_*j,t*_ }, as the *j*^*th*^ resample is obtained. Here *θ* is the mean of the standard exponential distribution;

1. a new *T ×* 2 sorting matrix *S*_2_ is defined and the *T* members of the set {*x*_*j,t*_} for the *j*^*th*^ resample obtained in Step 5 is reordered in an increasing order of magnitude and placed in column 1. The sorted *I*_*ord*_ values (Step 2) are placed in column 2 of *S*_2_;
2. matrix *S*_2_ is sorted according to the second column so that the order {1, 2, …, *T*} is there restored. The jointly sorted elements of column 1 is denoted by {*x*_*S,j,t*_}, where *S* recalls the sorting step;
3. Repeat Steps 1 to 7 a large number of times.

## 5. The application of the maximum entropy bootstrap

In what follows, the proposed procedure is presented in a step-by-step fashion.

1. For each time series 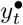 and 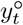 the bootstrap procedure is applied so that B= 100 “bona fide” replications are available, i.e. 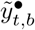 and 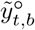;
2. for both the series, the row vector related to the last observation *T* is extracted, i.e.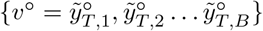 and 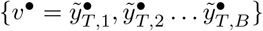
3. the expected values (𝔼(*v*^•^) 𝔼(*v*^°^)) are then extracted as well as the ≈ 95% confidence intervals (*CI*^•^ and *CI*^°^), computed according to the t–percentile method. The explanation of the T–percentile method goes beyond the scope of this paper, therefore the interested reader is referred to the excellent paper by Berkowitz and Kilian (2000).

In particular, the lower (upper) CIs will be the lower (upper) bounds of our estimator while the quantities 𝔼(*v*^•^) 𝔼(*v*^°^) are estimated through the mean operator, i.e.

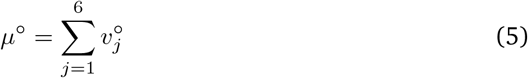

and

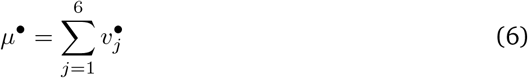

At this point, it is worth emphasizing that the procedure not only, as just seen, requires very little in terms of data but can be run in an automatic fashion. Once the data become available, one has just to divide them according to the subsets Ω i.e. Ω^*•*^ and the code will process the new data in an automatic way. The procedure is also very fast as the computing time needed for the generation of the bootstrap samples requires less than 2 minutes. Both code and data used for this Paper are freely made available for any researcher who would consider using it.

## 6. Empiricical evidences

In order to give the reader the opportunity to gain a better insight, in Figure 2 – 5 the time series of the variable *C* (see Eqn. 2) is reported for each region. Note that sudden variations (i.e. Bolzano in Figure 5, Valle D’Aosta in Figure 4 and Molise and CAmpania in Figure 3) are due to the little number of test administrated (denominator of the variable *C*_*T*_ (2))

**Fig. 1.**
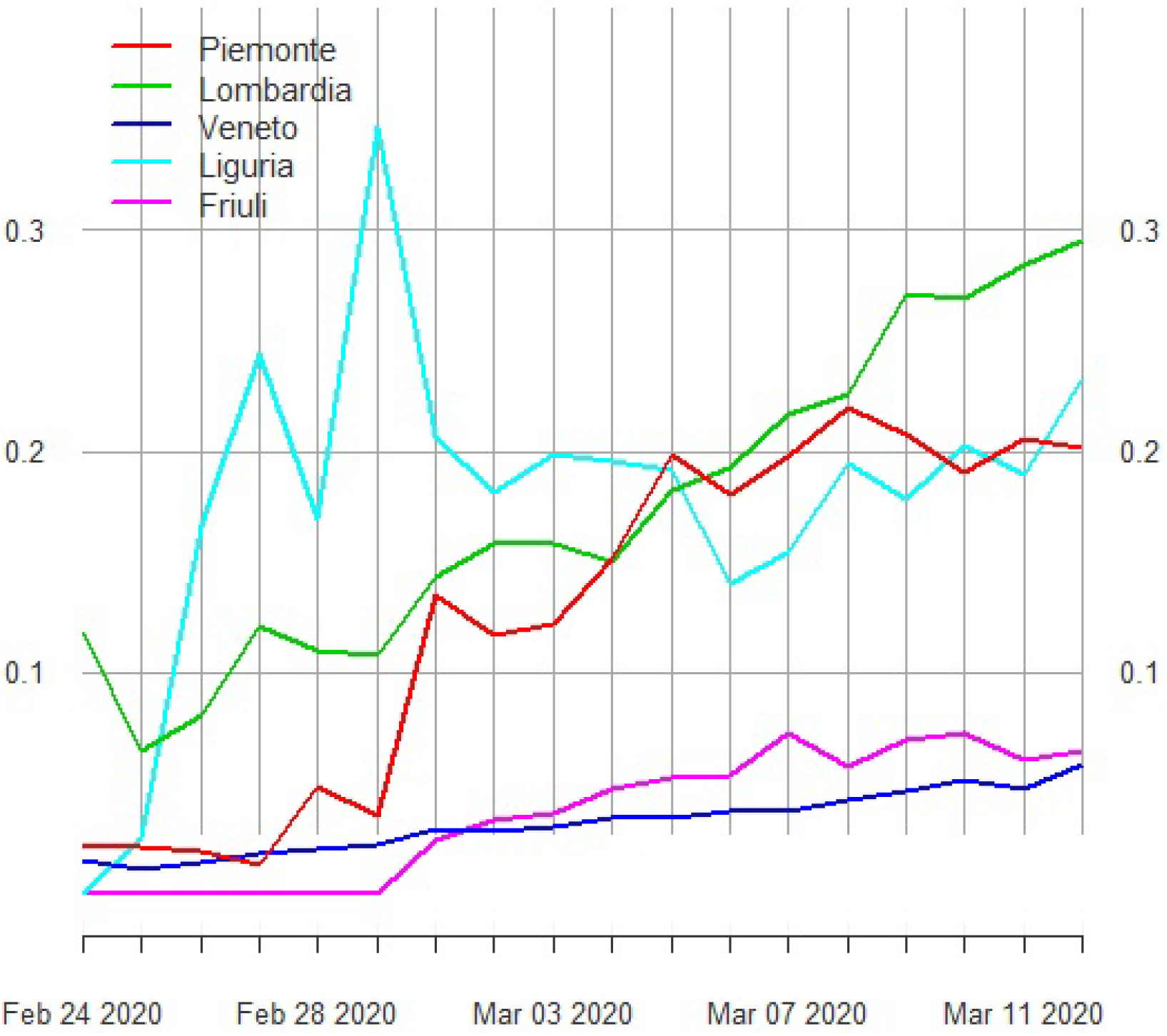
Percentage ratio deaths / new cases for the following Italian regions: Piemonte, Lombardia, Veneto, Liguria and Friuli-Venezia-Giulia

**Fig. 2.**
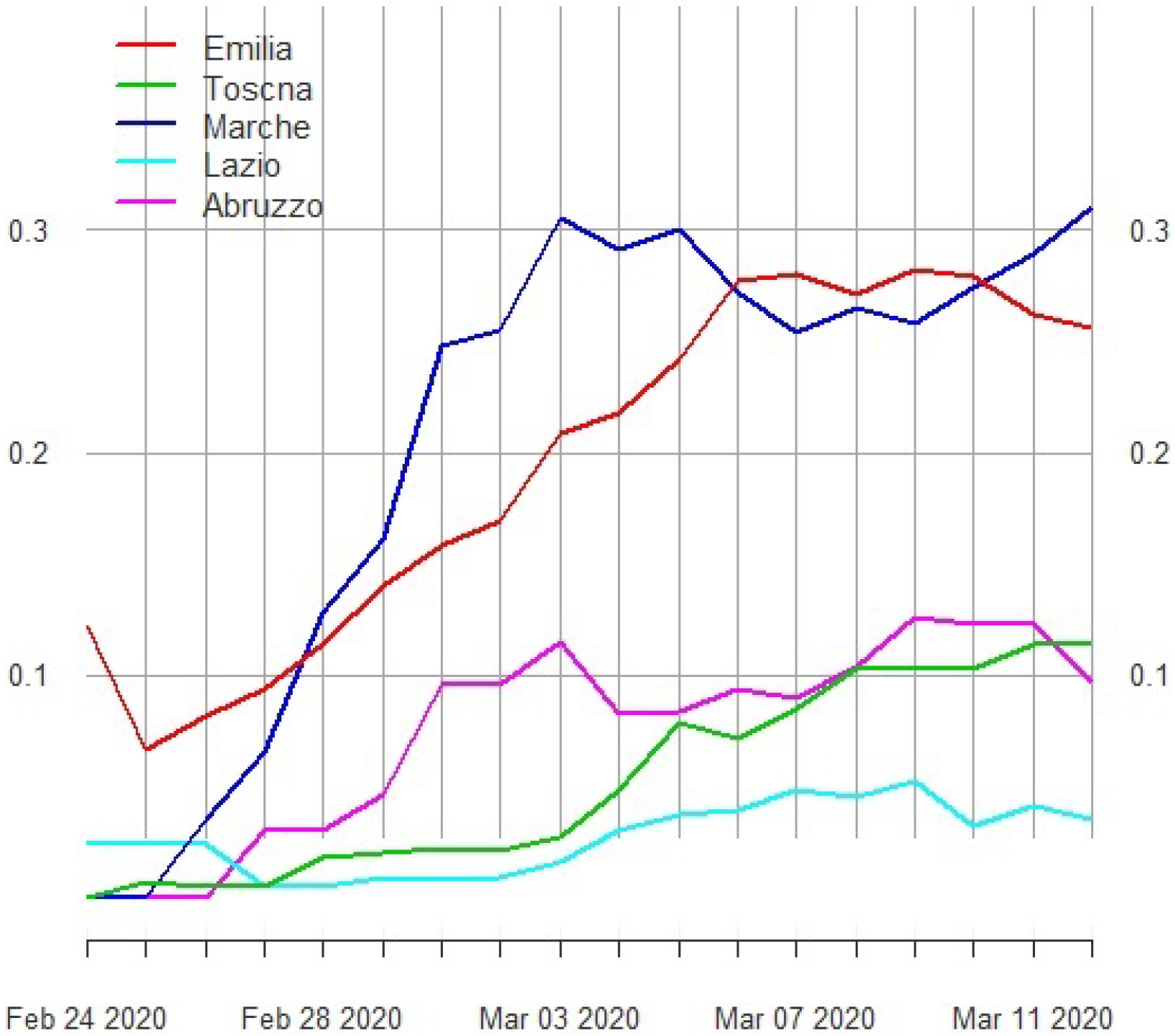
Percentage ratio deaths / new cases for the following Italian regions Emilia, Toscana, Marche, Lazio and Abruzzo

**Fig. 3.**
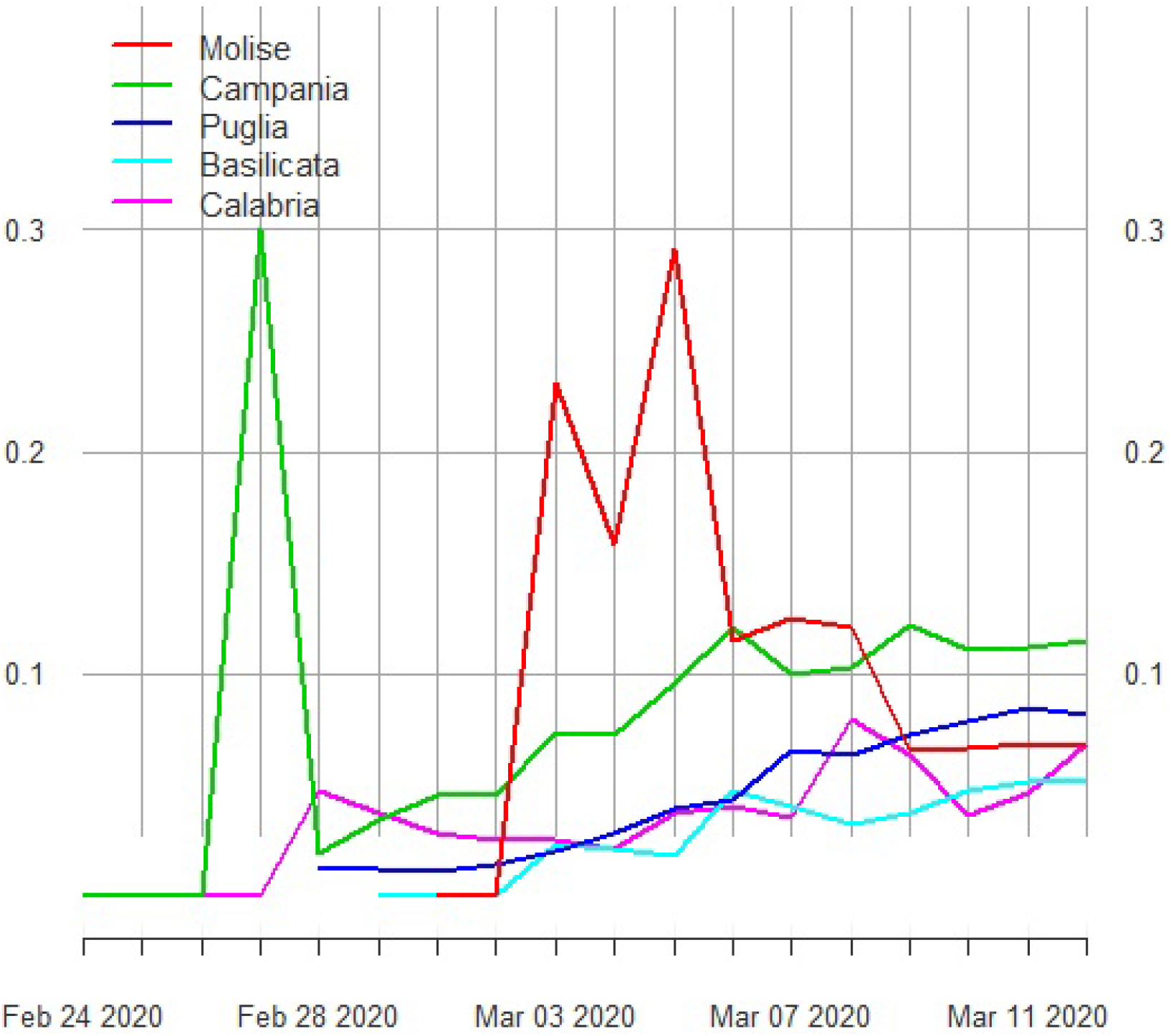
Percentage ratio deaths / new cases for the following Italian regions: Molise, Campania, Puglia, Basilicata and Calabria

**Fig. 4.**
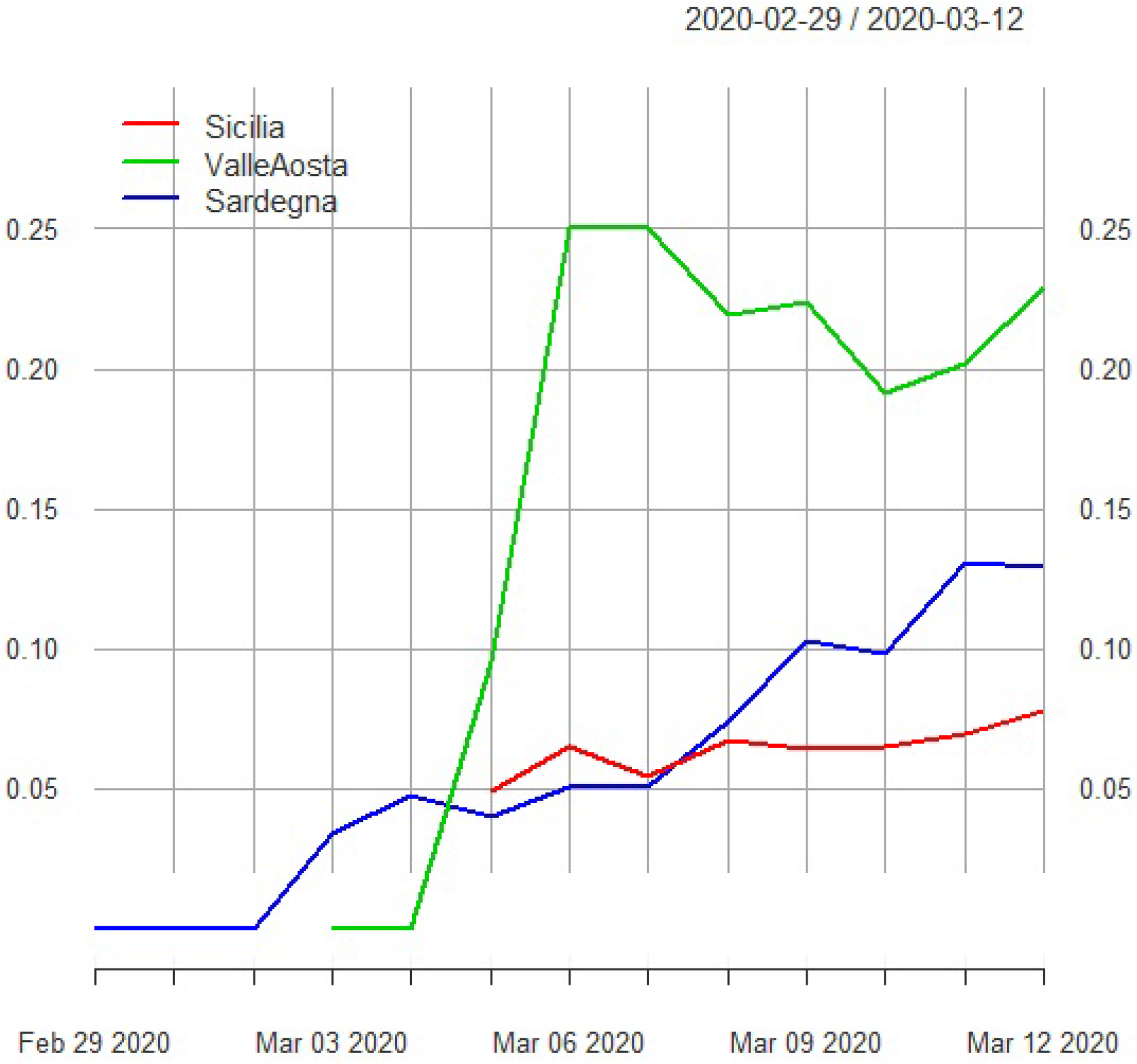
Percentage ratio deaths / new cases for the following Italian regions: Sicilia, Valle d’Aosta, Sardegna)

**Fig. 5.**
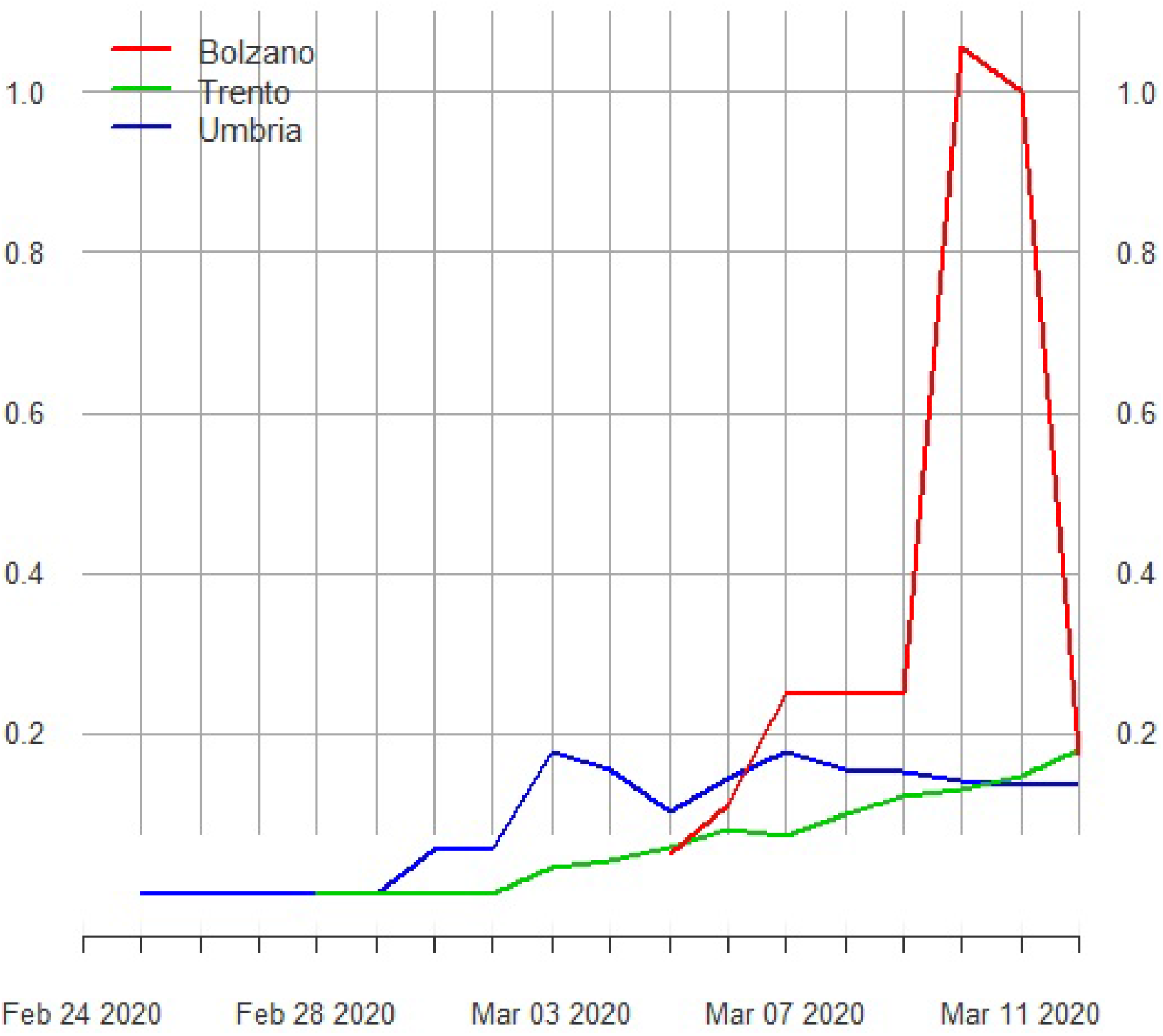
xxxxxxxxxxxxx

That said, the main result of the paper is summarized by Table 2, where three estimates of the number of infected people are reported by region. The regions belonging to the set Ω^°^ (i.e. no deaths) are in Italics (all the others belong to the set Ω^•^). In the column “Mean” and Lower (Upper) Bounds the bootstrap estimates computed according to Eqn 5 and 6 and the Lower (Upper) Bounds the lower (upper) bootstrap CIs are respectively reported. The column denominated “Official Cases” accounts for the number of official cases released by the Italian Authorities whereas the column “Morbidity” expresses the percentage ratio between *µ*^•^ (5) or *µ*^°^ (6) and the actual population of each region.

**Table 1.** Association found between the regions belonging to Ω^°^ and those in Ω^*•*^ according to the minimum distance *π*

| $\Omega^\circ$ | $\Omega^\bullet$ | $\pi$ |
| --- | --- | --- |
| Basilicata | Veneto | 0.0389 |
| Calabria | Campania | 0.6211 |
| Molise | Lazio | 0.4212 |
| Sardegna | Abruzzo | 0.0157 |
| Trento | Abruzzo | 0.00186 |
| Umbria | Sicilia | 0.01398 |

**Table 2.** Estimation of the number of people infected from CoViD–19 by Italian regions. Lower and Upper Bounds are computed through the Bootstrap t–percentile method whereas the mean values is computed as in (5) and (6) In Italics the regions belonging to the set Ω^°^ are reported

|  | Lower Bound | Mean | Upper Bound | Official Cases | Population | morbidity |
| --- | --- | --- | --- | --- | --- | --- |
| Abruzzo | 526 | 600 | 807 | 78 | 1.311.580 | 0,06 |
| *Basilicata* | 48 | 54 | 70 | 8 | 562.869 | 0,01 |
| Bolzano | 697 | 730 | 795 | 103 | 531.178 | 0,15 |
| *Calabria* | 182 | 238 | 493 | 32 | 1.947.131 | 0,03 |
| Campania | 988 | 1292 | 2676 | 174 | 5.801.692 | 0,05 |
| Emilia Romagna | 10980 | 12299 | 14897 | 1758 | 4.459.477 | 0,33 |
| Friuli Venezia Giulia | 983 | 1201 | 2514 | 148 | 1.215.220 | 0,21 |
| Lazio | 1485 | 1680 | 2089 | 172 | 5.879.082 | 0,04 |
| Liguria | 1346 | 1608 | 1995 | 243 | 1.550.640 | 0,13 |
| Lombardia | 37744 | 45020 | 49723 | 6896 | 10.060.574 | 0,49 |
| Marche | 3151 | 3891 | 4593 | 570 | 1.525.271 | 0,30 |
| *Molise* | 119 | 134 | 167 | 16 | 305.617 | 0,05 |
| Piemonte | 3216 | 3703 | 4217 | 554 | 4.356.406 | 0,10 |
| Puglia | 490 | 670 | 1292 | 98 | 4.029.053 | 0,03 |
| *Sardegna* | 244 | 278 | 375 | 39 | 1.639.591 | 0,02 |
| Sicilia | 776 | 865 | 1098 | 111 | 4.999.891 | 0,02 |
| Toscana | 2352 | 2755 | 3965 | 352 | 3.729.641 | 0,11 |
| *Trento* | 670 | 764 | 1028 | 102 | 541.098 | 0,19 |
| *Umbria* | 432 | 481 | 611 | 62 | 882.015 | 0,07 |
| Valle Aosta | 139 | 183 | 356 | 26 | 125.666 | 0,28 |
| Veneto | 8382 | 9343 | 12028 | 1297 | 4.905.854 | 0,25 |
| Totale Italia | 74.950 | 87.789 | 105.789 | 12.839 | 60359546 | 0,18 |

By examining the data for the whole Country, it is clear how the data collected by the Italian Authorities on the positive cases severely underestimate the current situation by a factor of about 8. As expected, the top three regions in terms of number of infected persons are Lombardia, Emilia Romagna and Veneto, where the estimated infected population is respectively (bootstrap mean) around 45,020, 12,299 and 9,343.

On the other hand, the risk of contagion is relatively low in regions – mostly located in the Southern part of Italy – and in the island of Sardegna.

Regarding the regions included in the subset Ω^°^, the application of the Piccolo distance (*π*) generated the associations reported in Table

## 7. Conclusions

It is widespread opinion in the scientific community that current official data on the diffusion of SARS-CoV-2, responsible of the correlated disease, COIVD-19,among population, are likely to suffer from a strong downward bias.

In this scenario, the aim of this instant paper is twofold: fist, it can compute realistic figures on the effective number of people infected with SARS-CoV-2 in Italy; second, it can provide a methodology, which improves current state of art and can be used to compute similar figures in other countries.

Following Pueyo 2020, this paper proposes a methodology which starts from Italian data considered restively certain, such as the number of deaths and the number of people tested positive to the virus, and due to this:

1. allows a population wide estimation of infected people and the computation of related confidence intervals;
2. extends Pueyo 2020 methodology to regions and areas where no deaths have been yet registered.

The entire procedure has been written in the programming language R and uses official data as published by the Italian Government. The whole code is made available upon request to any researcher who would consider using it.

Obtained results show that, while official data at March the 12th report 12.839 cases in Italy, people infected with the SARS-CoV-2 could be as high as 105.789. If this estimate were correct, mortality rates would decrease as its denominator increases, compared to what is calculated in official statistics.

On the other hand, considering that, in absence of strong actions, such as the decreasing of social distance among people and that the average doubling time for the Coronavirus (that is, the time it takes to double cases, on average) is 6.2 days (Pueyo (2020)), the pandemic is to be regarded as much more dangerous than currently foreseen.To overcome the crisis, international solidarity together wit, strong and coordinated actions among countries will be crucial. It is even worth to stress that at micro level everyone is called to act with the greatest responsibility, increasing social distance and respecting what imposed by authorities. Stay at home, and, if you can, do research on this topic, every contribution could be crucial.

## Data Availability

ALl the data employed are freely available on the Internet

## 8. Acknowledgments

The author is deeply grateful to Dr. Luigi Di Landro for the generous help in the proof-reading process.

## Notes

### Competing Interest Statement

The authors have declared no competing interest.

### Funding Statement

no funds

## References and links

Berkowitz, J., and Kilian, L. (2000), “Recent developments in bootstrapping time series,” Econometric Reviews, 19(1), 1–48.

Carlstein, E. et al. (1986), “The use of subseries values for estimating the variance of a general statistic from a stationary sequence,” The annals of statistics, 14(3), 1171–1179.

Koutris, A., Heracleous, M. S., and Spanos, A. (2008), “Testing for nonstationarity using maximum entropy resampling: A misspecification testing perspective,” Econometric Reviews, 27(4-6), 363–384.

Makridakis, S., and Hibon, M. (1997), “ARMA models and the Box–Jenkins methodology,” Journal of Forecasting, 16(3), 147–163.

Piccolo, D. (1990), “A distance measure for classifying ARIMA models,” Journal of Time Series Analysis, 11(2), 153–164.

Piccolo, D. (2007), Statistical issues on the AR metric in time series analysis,, in Proceedings of the SIS 2007 intermediate conference” Risk and Prediction, pp. 221–232.

Pueyo, T. (2020), Coronavirus: Why You Must Act Now,, in https://medium.com/@tomaspueyo/coronavirus-act-today-or-people-will-die-f4d3d9cd99ca.

Vinod, H. D., López-de Lacalle, J. et al. (2009), “Maximum entropy bootstrap for time series: the meboot R package,” Journal of Statistical Software, 29(5), 1–19.

